# Survival benefits of varying physical activity levels in a heterogeneous colorectal cancer cohort: The Disparities and Cancer Epidemiology (DANCE) study

**DOI:** 10.64898/2026.06.23.26356372

**Authors:** Sarah M Lima, Chiranjeev Dash, Jaeil Ahn, Rui Zhang, Stacy M Post, Siddhi Patil, Candy Promprasert, Batsirai Mabvakure, Reem Muhsen, Ann G Schwartz, Julie Ruterbusch, Angela S Wenzlaff, Mei-Chin Hseih, Elena M Stoffel, Kristen S Purrington, Laura S Rozek

## Abstract

**Background:** Recreational physical activity has been shown to improve survival among colorectal cancer (CRC) patients. With a growing survivor population, it is necessary to understand whether there are survival benefits across physical activity levels and across sociodemographic and clinical features.

**Methods:** Disparities and Cancer Epidemiology (DANCE) is a population-based cohort of CRC survivors from metro-Detroit and Louisiana. Self-reported moderate and vigorous recreational physical activity was modeled continuously and categorically as none, low (<7.5 MET-hrs/wk), and high (7.5+ MET-hrs/wk). Survival models estimated hazard ratios (HRs) for physical activity with all-cause and CRC-specific survival. Models were stratified by sociodemographic and clinical features; cross-product terms estimated interaction with physical activity.

**Results:** Of 1,107 participants, 26.5% were inactive, 49.1% had low physical activity, and 24.5% had high physical activity. Compared to inactivity, low activity was associated with 44% higher overall survival (HR= 0.56, 95% CI: 0.42, 0.75), and high activity with 66% higher survival (HR=0.34, 95% CI: 0.22, 0.53; P-trend=0.01). Adjustment for comorbidities, quality of life, BMI, and BMI-change did not alter results. Results remained significant for CRC-specific survival (low: HR=0.65, 95% CI: 0.44, 0.96; high: HR=0.45, 95% CI: 0.25, 0.80). Associations were consistent across sociodemographic and clinical features other than BMI and race; survival benefits were larger among White survivors.

**Conclusion:** Any recreational physical activity is associated with longer overall and CRC-specific survival, regardless of sociodemographic or clinical characteristics for the most part. Any physical activity may have survival benefits for CRC survivors, but meeting physical activity guidelines may have the greatest benefit.

## Background

Recreational physical activity (i.e., non-occupational) has been suggested as a secondary prevention strategy for colorectal cancer (CRC), with documented associations with longer survival.^1,2^ Compared to drug therapy, physical activity is relatively inexpensive and may be a useful tool to improve survival for CRC survivors (defined as individuals diagnosed with CRC)^3^, including low-income or under-insured survivors.^4^ The recent CHALLENGE randomized controlled trial reported 37% longer overall survival among stage II-III CRC patients who were randomized to a structured exercise intervention.^5^

However, it remains unclear whether the survival benefits of physical activity exist uniformly across various physical activity levels or across sociodemographic and clinical characteristics among CRC survivors. CHALLENGE trial participants reported having 11.5 metabolic equivalent tasks (MET)-hrs/wk at baseline,^5^ or roughly 115-230 minutes of vigorous-moderate activity. This level exceeds the Physical Activity Guidelines for Americans and American Cancer Society’s (ACS) recommendations of at least 75 minutes of weekly vigorous physical activity or 150 minutes of weekly moderate activity.^6,7^ US data suggest only a quarter of Americans meet these guidelines.^8^ Given the low levels of physical activity in the US population, understanding whether even modest amounts of physical activity are associated with improved survival among individuals with CRC is critical.

CRC is among the most common cancer sites and a leading cause of cancer death among men and women in the US.^9^ In 2026, CRC became the leading cause of cancer death for individuals diagnosed under 50.^10^ Survival statistics are even starker for certain subgroups. Patients diagnosed at distant stage, which represent nearly a quarter of all CRC cases, have a 16% five-year survival rate,^11^ while non-Hispanic Black patients have a 34% higher mortality rate than non-Hispanic White patients.^12^ Such survival outcomes further highlight the need to identify modifiable factors that influence CRC survival.

Studying physical activity in a heterogenous CRC survivor population will provide generalizable epidemiologic evidence, with important implications for clinical and survivorship care. We examined associations between varying levels of recreational physical activity with all-cause and CRC-specific survival in the Disparities and Cancer Epidemiology (DANCE) cohort and assessed relationships across sociodemographic and clinical features.

## Methods

### Study design & population

This study was conducted in the DANCE cohort, a population-based cohort of CRC survivors from metropolitan Detroit, Michigan and Louisiana. Cases were identified and sampled through the Metropolitan Detroit Cancer Surveillance System (MDCSS) and Louisiana Tumor Registry (LTR). Some DANCE cases were originally enrolled in the Detroit Research on Cancer Survivors study (n=528), an MDCSS-based cohort of Black cancer survivors.^13^ Eligibility criteria include invasive CRC, diagnosed at age 20-79 in 2013-2022, lived within MDCSS/LTR catchment areas, and self-identified as Black or non-Hispanic White. Informed consent was obtained from all participants and IRB approval was obtained at participating sites.

### Exposures

A modified International Physical Activity Questionnaire queried moderate and vigorous physical activity at baseline interview. Frequency and duration of self-reported moderate and vigorous activity was used to calculate MET-hrs/wk.^14^ Information on walking was not collected in questionnaires, thus physical activity measures refer only to moderate and vigorous activity. Physical activity was operationalized as: none, low (<7.5 MET-hrs/wk), and high (≥7.5 MET-hrs/wk). The high physical activity group corresponds with physical activity guidelines.^6,7^

### Outcome

The outcomes of interest were all-cause survival and CRC-specific survival. Information on diagnosis date, vital status, and cause of death were obtained through linkage to the MDCSS and LTR. CRC-specific deaths were identified based on primary cause of death ICD–10 codes C18-C20.

### Statistical analysis

Chi-square tests and ANOVAs tested differences by physical activity level for categorical and continuous variables, respectively. Kaplan-Meier curves evaluated overall and CRC-specific survival according to physical activity level. Overall survival curves by physical activity were additionally stratified according to cancer stage at diagnosis and race. Cox proportional hazards models estimated hazard ratios (HRs) associated with physical activity and overall survival and Fine-Gray competing risks models estimated HRs for CRC-specific survival. Physical activity was evaluated categorically and continuously per 10-MET-hrs/wk. The proportional hazards assumption was tested through visual evaluation of survival curves by physical activity level and evaluation of significant interaction with time in survival models.

Main models adjusted for age at diagnosis, study site, sex, race (White (ref), Black), education (<high school, high school/GED, some college or more (ref)), stage at diagnosis (local (ref), regional, distant, unknown), subsite (colon (ref), rectum), and smoking status (never (ref), former, current). Additional models included adjustment for comorbidities (0-1 (ref), 2-3, 4+) and Functional Assessment of Cancer Therapy-Colorectal (FACT-C) score to address potential confounding related to functional ability. BMI models adjusted for BMI at baseline and change in BMI from pre-diagnosis to baseline interview (average 29 months). BMI-change was operationalized as ± half-SD in continuous BMI-change and classified as increased, stable, or decreased. Finally, we ran models adjusting for time between diagnosis and baseline interview.

We stratified overall survival models to assess whether the relationship between physical activity level and survival differs by sociodemographic or clinical characteristics. We stratified models according to the following factors: age at onset, sex, race, current BMI, BMI-change, comorbidities, Area Deprivation Index (ADI),^15^ stage at diagnosis, tumor subsite, and time between diagnosis to baseline interview. Multiplicative interaction terms between physical activity level and stratifying factors were used to estimate the presence of statistically significant interaction. CRC-specific survival was stratified by stage at diagnosis but not by other factors due to too few CRC-specific events across strata.

Due to race-based differences in CRC survival, physical activity, and confounding factors, we ran the above sequence of models by race. Survival time was restricted to ≤115 months due to violated proportional hazards assumption among White cases. To determine whether there are threshold effects for physical activity and survival, we ran a cubic spline regression with 5 percentile-based knots to assess the non-linearity of physical activity then further tested physical activity per 5-MET-hrs/wk at the following thresholds: 0-10 MET-hrs/wk; 0-20 MET-hrs/wk; 0-30 MET-hrs/wk; >30 MET-hrs/wk. Analyses were performed using SAS 9.4 (Cary, NC); figures generated using R v4.6.0.

## Results

Of the 1,107 CRC survivors, 26.5% of participants were classified as inactive, 49.1% as low physical activity (<7.5 MET-hrs/wk), and 24.5% as high physical activity (7.5+ MET-hrs/wk; **Table 1**). We found that women, Black participants, participants with lower education, lower income, and greater area deprivation were more likely to be inactive. We also detected significant differences in physical activity by clinical factors, including current and pre-diagnostic BMI, comorbidities, FACT-C score, and stage at diagnosis, though we did not detect differences in BMI-change or distribution of subsite. Participants with high physical activity had better prognostic clinical profiles.

**Table 1.**
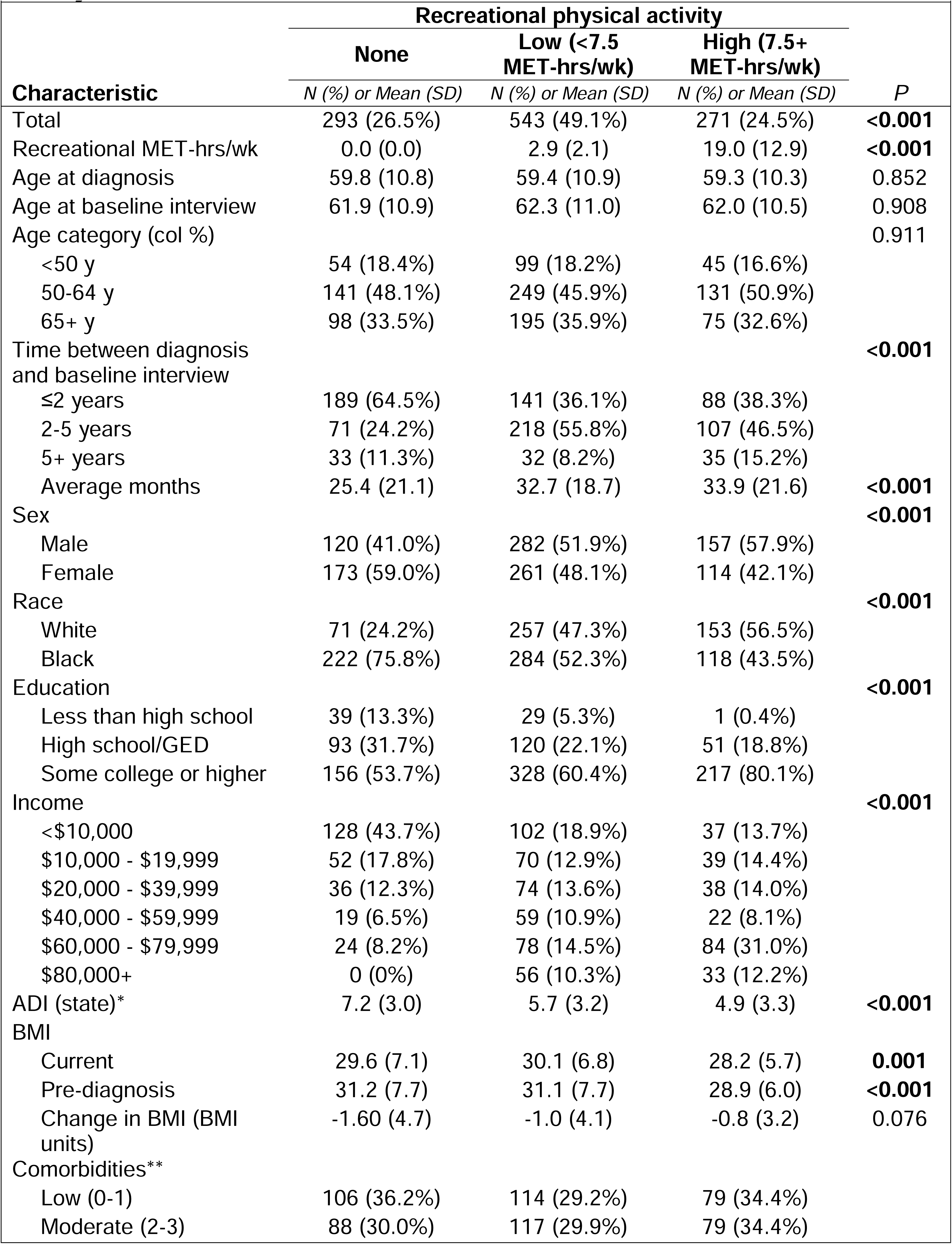
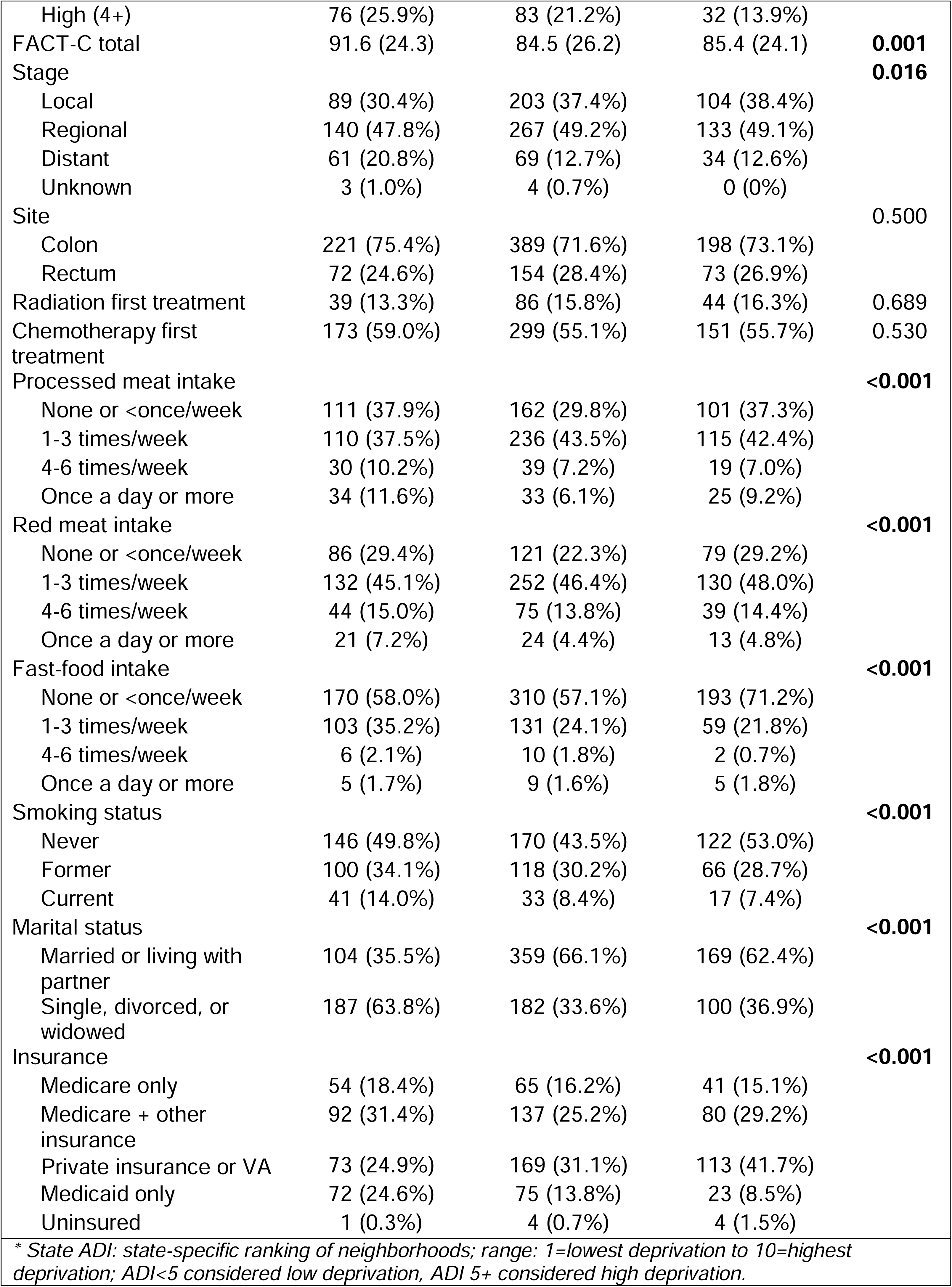
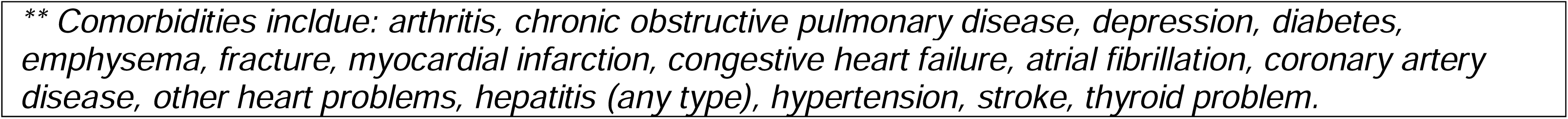
Descriptive statistics of DANCE (N=1107) by self-reported recreational physical activity.

| Characteristic | Recreational physical activity |  |  | P |
| --- | --- | --- | --- | --- |
|  | None | Low (<7.5 MET-hrs/wk) | High (7.5+ MET-hrs/wk) |  |
|  | N (%) or Mean (SD) | N (%) or Mean (SD) | N (%) or Mean (SD) |  |
| Total | 293 (26.5%) | 543 (49.1%) | 271 (24.5%) | **<0.001** |
| Recreational MET-hrs/wk | 0.0 (0.0) | 2.9 (2.1) | 19.0 (12.9) | **<0.001** |
| Age at diagnosis | 59.8 (10.8) | 59.4 (10.9) | 59.3 (10.3) | 0.852 |
| Age at baseline interview | 61.9 (10.9) | 62.3 (11.0) | 62.0 (10.5) | 0.908 |
| Age category (col %) |  |  |  | 0.911 |
| <50 y | 54 (18.4%) | 99 (18.2%) | 45 (16.6%) |  |
| 50-64 y | 141 (48.1%) | 249 (45.9%) | 131 (50.9%) |  |
| 65+ y | 98 (33.5%) | 195 (35.9%) | 75 (32.6%) |  |
| Time between diagnosis and baseline interview |  |  |  | **<0.001** |
| ≤2 years | 189 (64.5%) | 141 (36.1%) | 88 (38.3%) |  |
| 2-5 years | 71 (24.2%) | 218 (55.8%) | 107 (46.5%) |  |
| 5+ years | 33 (11.3%) | 32 (8.2%) | 35 (15.2%) |  |
| Average months | 25.4 (21.1) | 32.7 (18.7) | 33.9 (21.6) | **<0.001** |
| Sex |  |  |  | **<0.001** |
| Male | 120 (41.0%) | 282 (51.9%) | 157 (57.9%) |  |
| Female | 173 (59.0%) | 261 (48.1%) | 114 (42.1%) |  |
| Race |  |  |  | **<0.001** |
| White | 71 (24.2%) | 257 (47.3%) | 153 (56.5%) |  |
| Black | 222 (75.8%) | 284 (52.3%) | 118 (43.5%) |  |
| Education |  |  |  | **<0.001** |
| Less than high school | 39 (13.3%) | 29 (5.3%) | 1 (0.4%) |  |
| High school/GED | 93 (31.7%) | 120 (22.1%) | 51 (18.8%) |  |
| Some college or higher | 156 (53.7%) | 328 (60.4%) | 217 (80.1%) |  |
| Income |  |  |  | **<0.001** |
| <\$10,000 | 128 (43.7%) | 102 (18.9%) | 37 (13.7%) | |
| \$10,000 - \$19,999 | 52 (17.8%) | 70 (12.9%) | 39 (14.4%) | |
| \$20,000 - \$39,999 | 36 (12.3%) | 74 (13.6%) | 38 (14.0%) | |
| \$40,000 - \$59,999 | 19 (6.5%) | 59 (10.9%) | 22 (8.1%) | |
| \$60,000 - \$79,999 | 24 (8.2%) | 78 (14.5%) | 84 (31.0%) | |
| \$80,000+ | 0 (0%) | 56 (10.3%) | 33 (12.2%) | |
| ADI (state)* | 7.2 (3.0) | 5.7 (3.2) | 4.9 (3.3) | **<0.001** |
| BMI |  |  |  |  |
| Current | 29.6 (7.1) | 30.1 (6.8) | 28.2 (5.7) | **0.001** |
| Pre-diagnosis | 31.2 (7.7) | 31.1 (7.7) | 28.9 (6.0) | **<0.001** |
| Change in BMI (BMI units) | -1.60 (4.7) | -1.0 (4.1) | -0.8 (3.2) | 0.076 |
| Comorbidities** |  |  |  |  |
| Low (0-1) | 106 (36.2%) | 114 (29.2%) | 79 (34.4%) |  |
| Moderate (2-3) | 88 (30.0%) | 117 (29.9%) | 79 (34.4%) |  |
| High (4+) | 76 (25.9%) | 83 (21.2%) | 32 (13.9%) |  |
| FACT-C total | 91.6 (24.3) | 84.5 (26.2) | 85.4 (24.1) | **0.001** |
| Stage |  |  |  | **0.016** |
| Local | 89 (30.4%) | 203 (37.4%) | 104 (38.4%) |  |
| Regional | 140 (47.8%) | 267 (49.2%) | 133 (49.1%) |  |
| Distant | 61 (20.8%) | 69 (12.7%) | 34 (12.6%) |  |
| Unknown | 3 (1.0%) | 4 (0.7%) | 0 (0%) |  |
| Site |  |  |  | 0.500 |
| Colon | 221 (75.4%) | 389 (71.6%) | 198 (73.1%) |  |
| Rectum | 72 (24.6%) | 154 (28.4%) | 73 (26.9%) |  |
| Radiation first treatment | 39 (13.3%) | 86 (15.8%) | 44 (16.3%) | 0.689 |
| Chemotherapy first treatment | 173 (59.0%) | 299 (55.1%) | 151 (55.7%) | 0.530 |
| Processed meat intake |  |  |  | **<0.001** |
| None or <once/week | 111 (37.9%) | 162 (29.8%) | 101 (37.3%) |  |
| 1-3 times/week | 110 (37.5%) | 236 (43.5%) | 115 (42.4%) |  |
| 4-6 times/week | 30 (10.2%) | 39 (7.2%) | 19 (7.0%) |  |
| Once a day or more | 34 (11.6%) | 33 (6.1%) | 25 (9.2%) |  |
| Red meat intake |  |  |  | **<0.001** |
| None or <once/week | 86 (29.4%) | 121 (22.3%) | 79 (29.2%) |  |
| 1-3 times/week | 132 (45.1%) | 252 (46.4%) | 130 (48.0%) |  |
| 4-6 times/week | 44 (15.0%) | 75 (13.8%) | 39 (14.4%) |  |
| Once a day or more | 21 (7.2%) | 24 (4.4%) | 13 (4.8%) |  |
| Fast-food intake |  |  |  | **<0.001** |
| None or <once/week | 170 (58.0%) | 310 (57.1%) | 193 (71.2%) |  |
| 1-3 times/week | 103 (35.2%) | 131 (24.1%) | 59 (21.8%) |  |
| 4-6 times/week | 6 (2.1%) | 10 (1.8%) | 2 (0.7%) |  |
| Once a day or more | 5 (1.7%) | 9 (1.6%) | 5 (1.8%) |  |
| Smoking status |  |  |  | **<0.001** |
| Never | 146 (49.8%) | 170 (43.5%) | 122 (53.0%) |  |
| Former | 100 (34.1%) | 118 (30.2%) | 66 (28.7%) |  |
| Current | 41 (14.0%) | 33 (8.4%) | 17 (7.4%) |  |
| Marital status |  |  |  | **<0.001** |
| Married or living with partner | 104 (35.5%) | 359 (66.1%) | 169 (62.4%) |  |
| Single, divorced, or widowed | 187 (63.8%) | 182 (33.6%) | 100 (36.9%) |  |
| Insurance |  |  |  | **<0.001** |
| Medicare only | 54 (18.4%) | 65 (16.2%) | 41 (15.1%) |  |
| Medicare + other insurance | 92 (31.4%) | 137 (25.2%) | 80 (29.2%) |  |
| Private insurance or VA | 73 (24.9%) | 169 (31.1%) | 113 (41.7%) |  |
| Medicaid only | 72 (24.6%) | 75 (13.8%) | 23 (8.5%) |  |
| Uninsured | 1 (0.3%) | 4 (0.7%) | 4 (1.5%) |  |

\* State ADI: state-specific ranking of neighborhoods; range: 1=lowest deprivation to 10=highest deprivation; ADI<5 considered low deprivation, ADI 5+ considered high deprivation.
*\*\* Comorbidities include: arthritis, chronic obstructive pulmonary disease, depression, diabetes, emphysema, fracture, myocardial infarction, congestive heart failure, atrial fibrillation, coronary artery disease, other heart problems, hepatitis (any type), hypertension, stroke, thyroid problem.*

We found significant differences in survival probability by physical activity levels for all-cause and CRC-specific survival, with higher physical activity levels showing progressively higher survival (**Figure 1**). Overall survival probability was higher with higher activity levels regardless of stage at diagnosis (**Figure S1**) and race (**Figure S2**).

**Figure 1.**
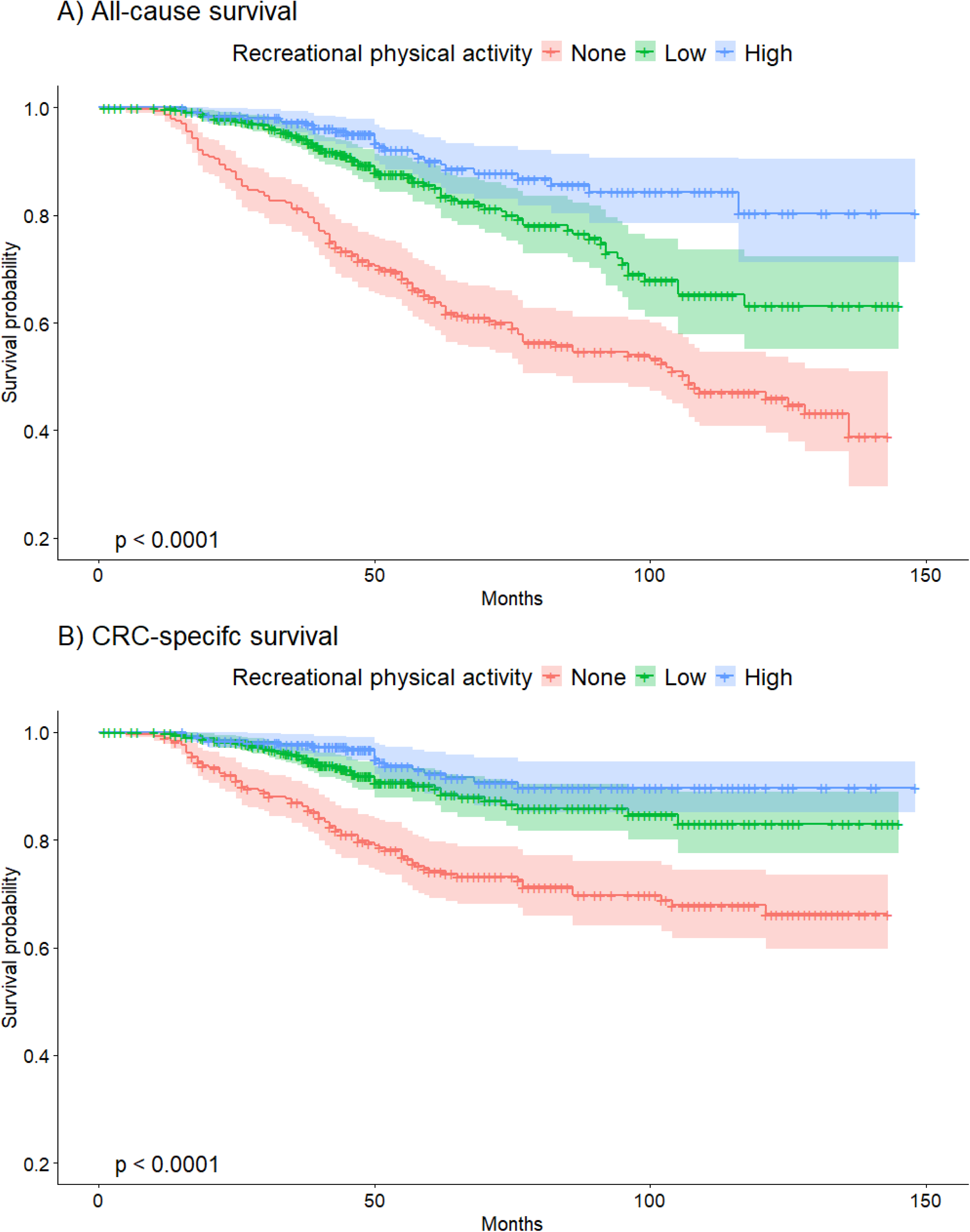
Survival probability of CRC survivors by recreational physical activity level for (A) all-cause survival and (B) CRC-specific survival.

**Table 2** reports associations for recreational physical activity with all-cause and CRC-specific survival. In main models, we found overall survival was 44% higher with low physical activity (HR= 0.56, 95% CI: 0.42, 0.75) and 66% higher with high physical activity (HR=0.34, 95% CI: 0.22, 0.53) compared to inactivity. We found significant trends with physical activity, where HR=0.73 (95% CI: 0.57, 0.93) per 10 MET-hrs/wk increase (P-trend =0.012). Associations were largely unchanged with further adjustment for comorbidities, FACT-C score, BMI, BMI-change, or time between diagnosis and baseline interview. Associations were attenuated when evaluating CRC-specific survival. We found longer CRC-specific survival associated with low physical activity (HR=0.65, 95% CI: 0.44, 0.96) and high physical activity (HR=0.45, 95% CI: 0.25, 0.80) compared to inactivity, though we did not detect a significant trend (P-trend=0.437). Adjustment for comorbidities, FACT-C score, and BMI change attenuated results such that associations for low activity and high activity were borderline nonsignificant. High physical activity showed a benefit for CRC-specific survival for both early-stage (HR=0.47, 95% CI: 0.20, 1.08) and late-stage cases (HR=0.47, 95% CI: 0.20, 1.07), whereas low physical activity did not show benefit for late-stage and only borderline benefit for early-stage cases (HR=0.57, 95% CI: 0.30, 1.07).

**Table 2.**
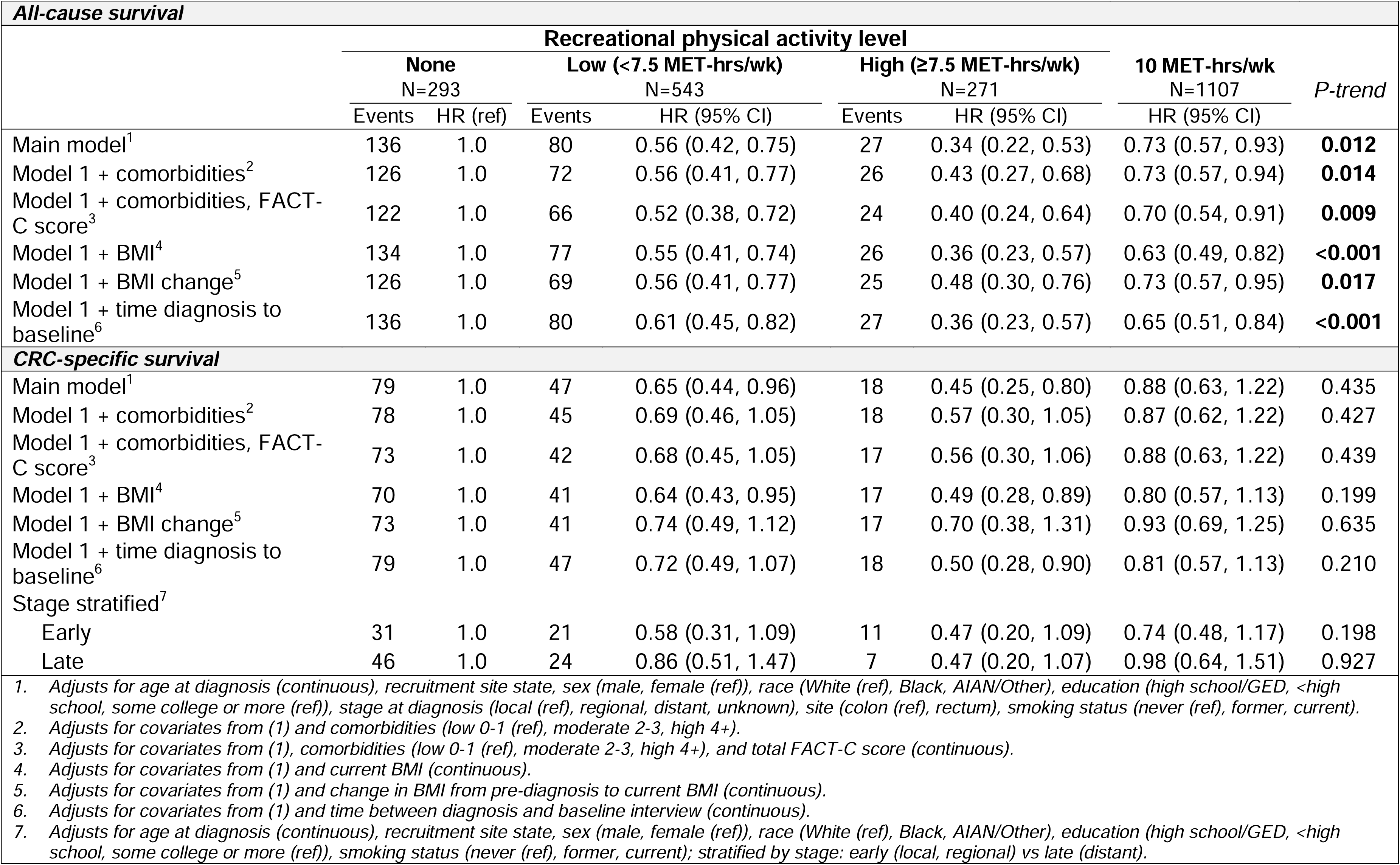
Recreational physical activity and all-cause and CRC-specific survival.

Results from our cubic spline regression indicated a threshold effect for physical activity at 30 MET-hrs/wk. In further testing physical activity at various MET-hr/wk thresholds (**Table S1**), we found a larger effect size per 5 MET-hrs/wk increase in activity for up to 30 MET-hrs/wk (HR_all-cause_=0.70, 95% CI: 0.59, 0.82) compared to assessing all levels of physical activity (HR_all-cause_=0.80, 95% CI: 0.70, 0.90). We found similar results with CRC-specific survival. Further, we detected a significantly higher risk of CRC death associated with physical activity when evaluating among survivors with >30 MET-hrs/wk.

Stratified results are reported in **Figure 2** according to low and high physical activity level. Compared to inactivity, low physical activity was associated with higher all-cause survival across sociodemographic and clinical characteristics. We found evidence of heterogenic effects by race (P-interaction = 0.012), in which low physical activity had a larger effect among White participants (HR=0.40, 95% CI: 0.22, 0.73) compared to Black participants (HR=0.60, 95% CI: 0.43, 0.85). Of note, the benefit of low physical activity was not statistically significant among participants with early-onset CRC, late-stage diagnosis, lean BMI (18.5-24.9), had a BMI-increase, or lived in high-deprivation neighborhood (ADI ≥5). Effect sizes for high physical activity were larger than low activity, but overall findings were generally consistent. We detected a significant interaction between high activity and current BMI, where physical activity had null associations among lean participants but a progressively larger effect with overweight and obesity. We did not find significant associations between high physical activity and overall survival among participants diagnosed with early-onset CRC or rectal cancer, with stable or increased BMI, or with 2-3 comorbidities. Effect sizes for both low and high physical were similar when evaluated according to time between diagnosis and baseline interview.

**Figure 2.**
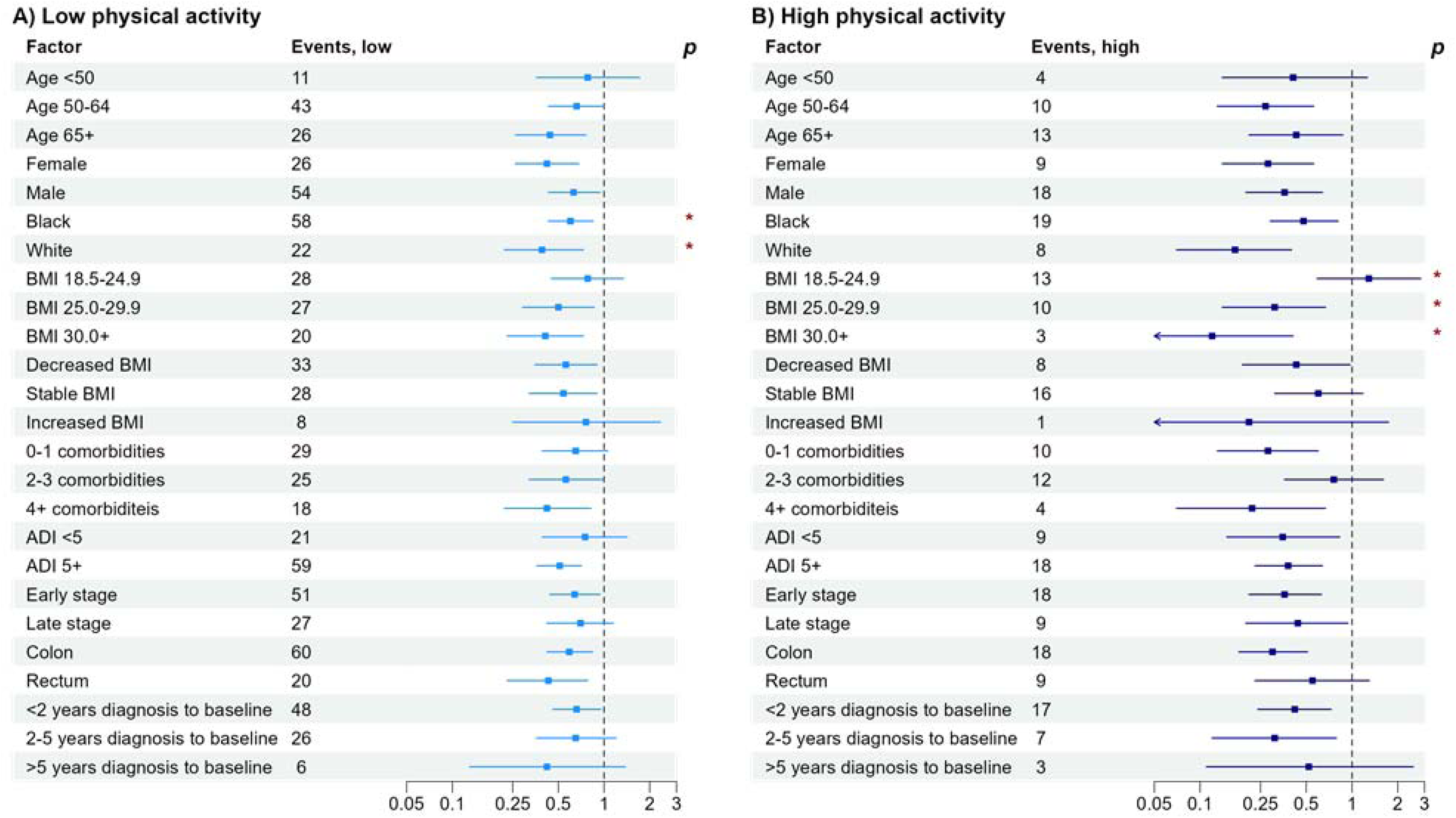
Stratified associations between physical activity and overall survival for (A) low physical activity and (B) high physical activity. P reports significant interaction, * = p<0.05. Each point represents the HR for physical activity level compared to inactivity in the specific subgroup.

**Figure 3** shows associations between physical activity level and overall survival according to race. Consistent with our overall models, we found a progressively higher survival benefit associated with low and high physical activity among both Black and White participants, though effect sizes were larger among White participants. As with previous models, adjustment for comorbidities, FACT-C, or BMI factors did not influence results. We did not find significant survival benefits for low or high activity among early-onset cases among either Black or White survivors. Among Black participants, physical activity showed survival benefits for survivors living in neighborhoods with ADI ≥5 (high deprivation) and lower household income categories, but we did find survival benefits for those living in neighborhoods with ADI <5 (low deprivation) or with household incomes ≥$60,000. In contrast, high activity was associated with longer survival among White participants regardless of ADI.

**Figure 3.**
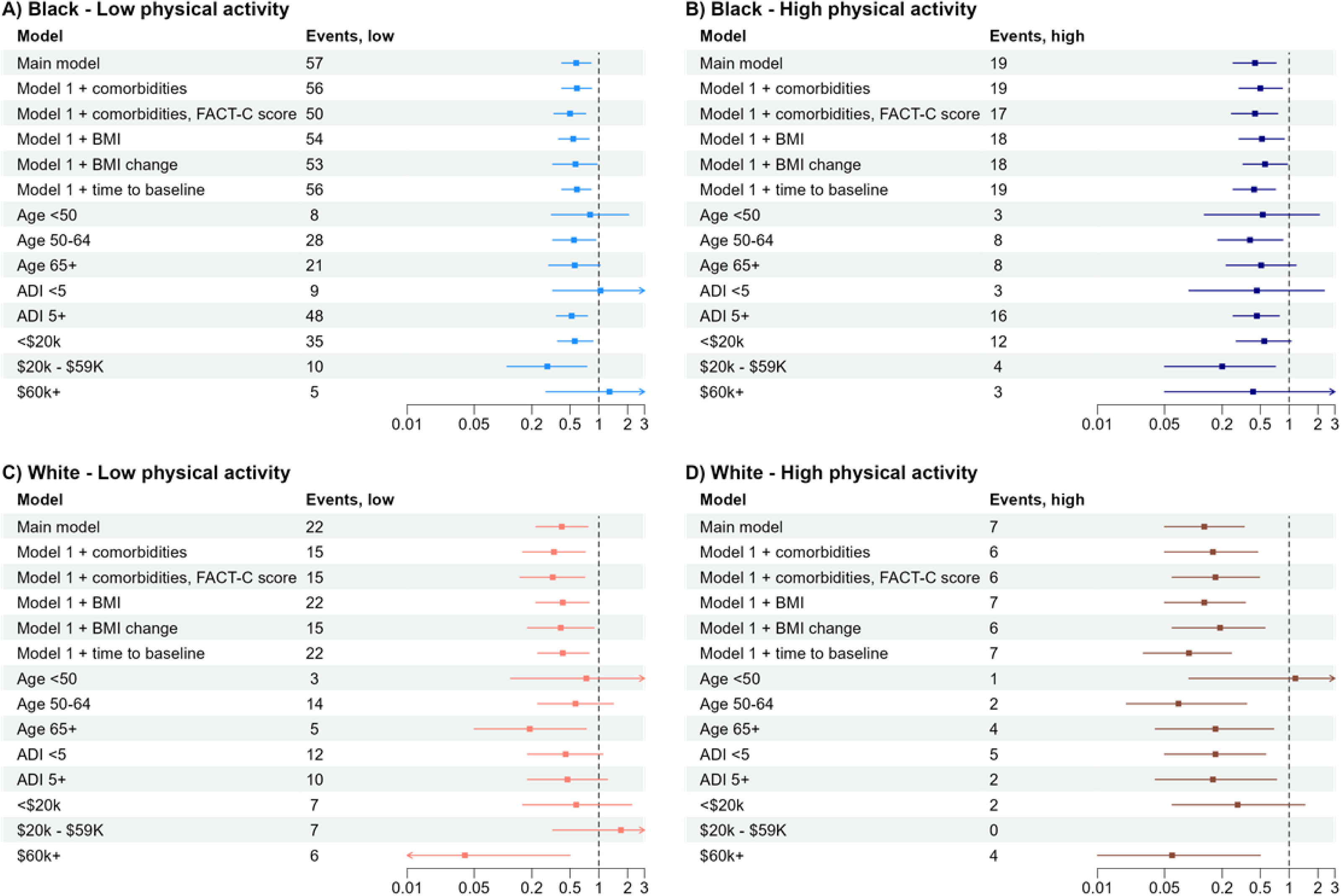
Race-specific associations between physical activity and overall survival. (A) Low physical activity among Black participants, (B) high physical activity among Black participants, (C) low physical activity among White participants, (D) high physical activity among White participants.

## Discussion

In this diverse CRC survivor cohort, we found moderate-vigorous recreational physical activity was associated with significantly longer overall survival and CRC-specific survival. Importantly, these data suggest that for CRC survivors, even modest amounts of physical activity have survival benefits, but that survival benefit is highest when meeting recommended physical activity levels.^6,7^ Thus, clinicians advising CRC survivors on lifestyle behaviors may wish to focus on the benefits of physical activity in any amount, though more (up to a threshold) appears to have greater benefits.

Our findings are consistent with results from numerous observational studies showing a CRC survival benefit from physical activity and results from the CHALLENGE trial.^5,16–18^ However, the CHALLENGE trial participants reported pre-intervention physical activity levels higher than guideline-recommended minimum activity levels (11 MET-hrs/wk vs 7.5 MET-hrs/wk), which only a quarter of Americans are estimated to meet.^6,8^ We show evidence that CRC survivors can benefit from physical activity even if they do not meet guidelines. Our results support findings that suggest small amounts of physical activity, including even 10 minutes of brisk walking, have significant survival benefits.^19,20^ Although current physical activity guidelines for the general population state that some physical activity is better than none,^6^ ACS guidelines for cancer survivors align with the US guidelines of 150-300 minutes of moderate intensity or 75-150 minutes of high intensity exercise per week.^2,7^ This puts CRC survivors at risk for “all-or-nothing thinking” with exercise: feeling like a failure for not meeting an exercise goal, and therefore not engaging in any exercise as a result.^21^ With a growing CRC survivor population, these results collectively indicate clinical messaging to survivors should focus any level of movement for prolonged survival.

Though not our primary objective, our threshold analysis suggested survival benefits of physical activity may peak at 30 MET-hrs/wk (equivalent to 600 minutes of moderate activity or 300 minutes of vigorous activity). Above this, physical activity may not have survival benefits and may even increase mortality risk, which we detected with CRC-specific survival. While we caution interpretation of these results given our limited power to examine associations among survivors with ≥30 MET-hrs/wk, these results align with preliminary reporting of relatively high prevalence of advanced adenomas among endurance runners. Given extreme physical activity can result in inflammation, ischemic injury, and other CRC-relevant pathways,^22,23^ further research is needed on the relationship between extreme physical activity levels and CRC endpoints.

Despite variation in physical activity by participant characteristics, we found the survival benefit largely does not differ by sociodemographic or clinical features. Notable exceptions where we did not observe a survival benefit across physical activity levels include individuals with early-onset CRC, lean BMI, and a BMI-increase from pre-diagnosis to baseline interview. These results may be reflective of underlying non-cancer mortality risk found in older CRC survivors and those with higher weight status. Physical activity has been shown to reduce the risk of cardiovascular-related death among cancer survivors, but cardiovascular mortality risk is relatively low for those age <50.^24,25^ Our finding of a protective effect among overweight and obese survivors but not survivors with lean BMI is in contrast to a prior study on physical activity and BMI combinations.^26^ While the CRC survivors with lean BMI at baseline may have experienced cancer or treatment related weight-loss, we nonetheless detected a protective effect with physical activity among those who experienced a BMI-decrease, adding further complexity to the relationship between BMI and mortality.^27,28^ Benefits of physical activity may be most prominent for survivors that have overweight or obesity but who did not gain weight.

Our results suggest the survival benefits of physical activity may differ by race, where the benefit may be larger among White survivors than Black survivors. Physical activity levels varied greatly by race and income, where over half of inactive participants have an income <$20,000 and 75% are Black. Importantly, we found physical activity had significant survival benefits among socioeconomically disadvantaged Black survivors, whether individual-level or neighborhood-level disadvantage. In contrast, Black survivors with high socioeconomic status (individual-level or neighborhood-level) did not see survival benefits from physical activity, while White survivors did regardless of socioeconomic factors and despite having less statistical power. These data require important public health considerations. While results suggest physical activity can improve survival for Black, socioeconomically disadvantaged CRC survivors—a group with low survival rates^12^—physical activity in these subgroups is low. Several studies have noted a lack of access to recreational physical activity resources, including public spaces such as sidewalks, parks, or recreational courts, that impact health outcomes in high-deprivation communities.^29,30^ Our null results among Black survivors with high socioeconomic status, while surprising, is supportive of the diminishing returns theory, which states that Black Americans see fewer health benefits from socioeconomic gains than White Americans.^31^ This may be explained by higher chronic stress and allostatic load stemming from interpersonal racial discrimination or other forms of intersectional stigma.^32–34^

A major strength of this study is the diversity and size of the DANCE CRC cohort. This enabled us to conduct analyses on varying physical activity levels and multiple stratified analyses, including race-specific models with additional stratification. We were also able to examine associations between physical activity and survival according to socioeconomic factors at the individual and neighborhood level. Our use of cohort data and multiple stratified analyses improves generalizability, revealing physical activity is largely beneficial to survival regardless of sociodemographic factors or clinical features. We also evaluated varying levels of physical activity, including operationalizing physical activity continuously to test for trends and categorically into clinically-actionable groups. This was further strengthened by assessing threshold effects of physical activity with all-cause and CRC-specific survival. Survey data allowed us to adjust for multiple important confounders, including comorbidities, FACT-C score for quality of life (including physical and functional quality of life), baseline BMI, and change in BMI from pre-diagnosis to baseline. Results were robust across these analyses and covariates. Furthermore, we did not detect interaction between physical activity and time between diagnosis and baseline interview, additional evidence that our results are not solely proxying disease severity or functional capacity to engage in exercise.

Limitations exist and should be noted. Although multiple models were used to address confounding from comorbidities, physical function, or advanced disease, residual confounding is possible and could lead to reverse causation. For example, individuals diagnosed with advanced CRC are at increased mortality risk and undergo intense therapies that may reduce energy or ability to engage in physical activity, both due to severity of disease. However, because we found larger effects for high physical activity compared to low physical activity and significant trends with MET-hrs/wk, we believe this bias is relatively limited. DANCE does not collect data on brisk walking, thus physical activity levels may be may be underestimated. Data on physical activity and comorbidities rely on self-report, thus are vulnerable to misclassification from social desirability or recall bias. While our cohort includes >1,100 participants, we were underpowered to conduct stratified analyses for CRC-specific survival. Finally, while we evaluated thresholds of physical activity as a sensitivity analysis, we had limited sample size for individuals with ≥30 MET-hr/wk and are underpowered to thoroughly evaluate threshold effects.

## Conclusion

Physical activity is associated with significantly longer survival for individuals with CRC, including modest physical activity, with an even larger benefit for individuals achieving guideline-recommended physical activity levels. Physical activity is associated with longer survival across demographic, socioeconomic, and clinical factors. Clinicians should emphasize the benefits of any physical activity to improve outcomes among CRC survivors.

## Data availability

The data underlying this article cannot be shared publicly due to privacy of individuals that participated in the study. The data will be shared on reasonable request to the corresponding author.

