## Supplemental materials for "Survival benefits of varying physical activity levels in a heterogeneous colorectal cancer cohort: The Disparities and Cancer Epidemiology (DANCE) study"

| **Table S1. METs threshold effects for all-cause and CRC-specific survival** | | | |
| --- | --- | --- | --- |
| ***All-cause survival*** | | | |
|  |  | **5 MET-hrs/wk** | *P-trend* |
|  |  | N=1107 |  |
| Model | Events | HR (95% CI) |  |
| METs modeled continuously, assuming linearity | 243 | 0.80 (0.70, 0.90) | **<0.001** |
| METs modeled continuously, testing thresholds |  |  |  |
| ≤10 METs threshold | 221 | 0.53 (0.38, 0.74) | **<0.001** |
| ≤20 METs threshold | 235 | 0.67 (0.56, 0.82) | **<0.001** |
| ≤30 METs threshold | 238 | 0.70 (0.59, 0.82) | **<0.001** |
| >30 METs threshold | 5 | * | * |
| ***Colorectal cancer-specific survival*** | | | |
|  |  | **5 MET-hrs/wk** |  |
|  |  | N=1107 | *P-trend* |
| Model | Events | HR (95% CI) |  |
| METS modeled continuously, assuming linearity | 144 | 0.88 (0.73, 1.05) | 0.153 |
| METs modeled continuously, testing thresholds |  |  |  |
| ≤10 METs threshold | 130 | 0.64 (0.43, 0.97) | **0.035** |
| ≤20 METs threshold | 139 | 0.76 (0.59, 0.98) | **0.034** |
| ≤30 METs threshold | 141 | 0.77 (0.62, 0.96) | **0.017** |
| >30 METs threshold | 3 | 4.59 (2.63, 8.02) | **<0.001** |
| *Adjusts for age at diagnosis (continuous), recruitment site state, sex (male, female (ref)), race (White (ref), Black, AIAN/Other), education (high school/GED, <high school, some college or more (ref)), stage at diagnosis (local (ref), regional, distant, unknown), site (colon (ref), rectum), smoking status (never (ref), former, current).*  ** Model unable to converge.* | | | |

**Figure S1.** **Survival probability according to recreational physical activity level stratified by stage at diagnosis.**

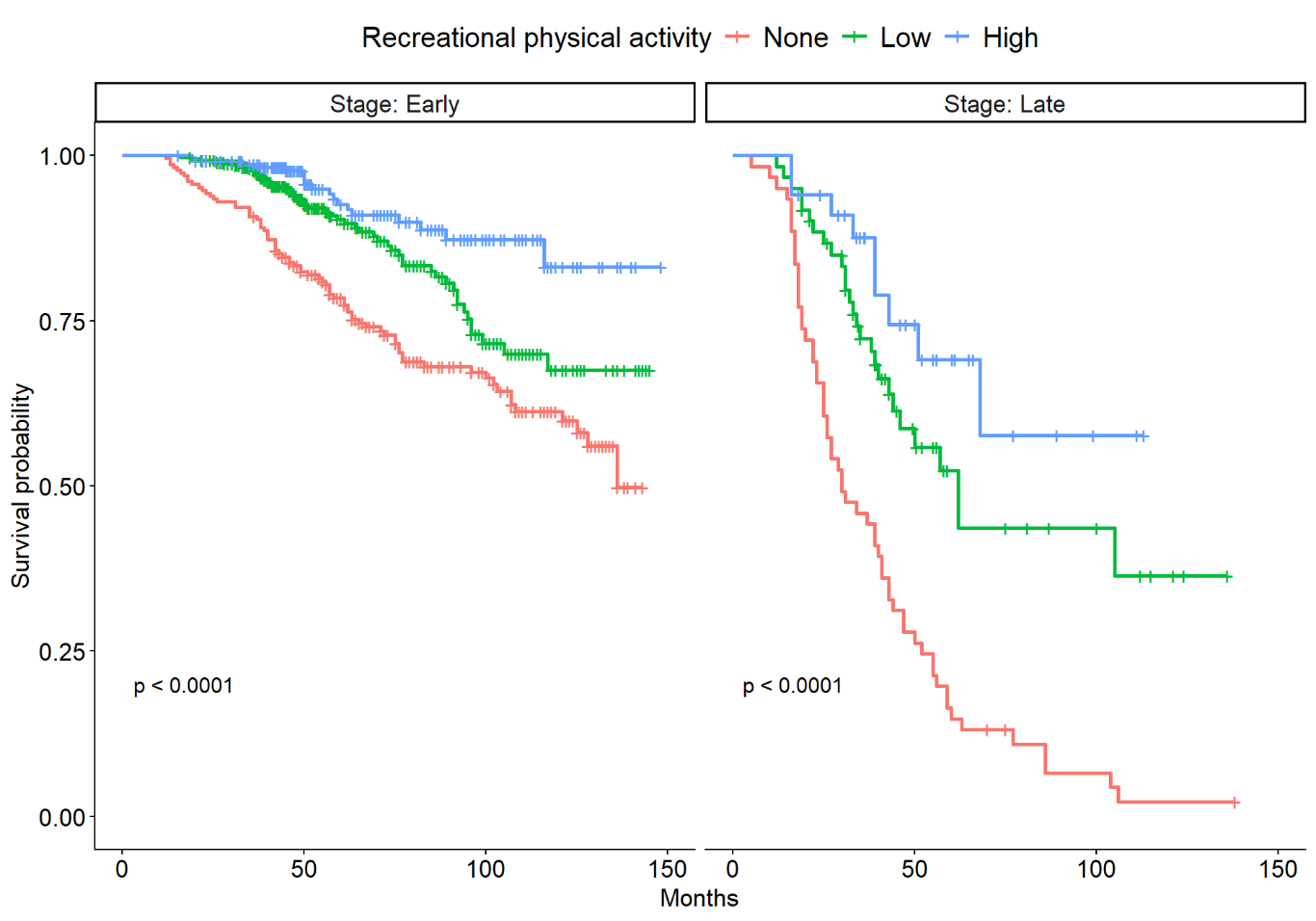

**Figure S2.** **Survival probability according to recreational physical activity level stratified by race.**

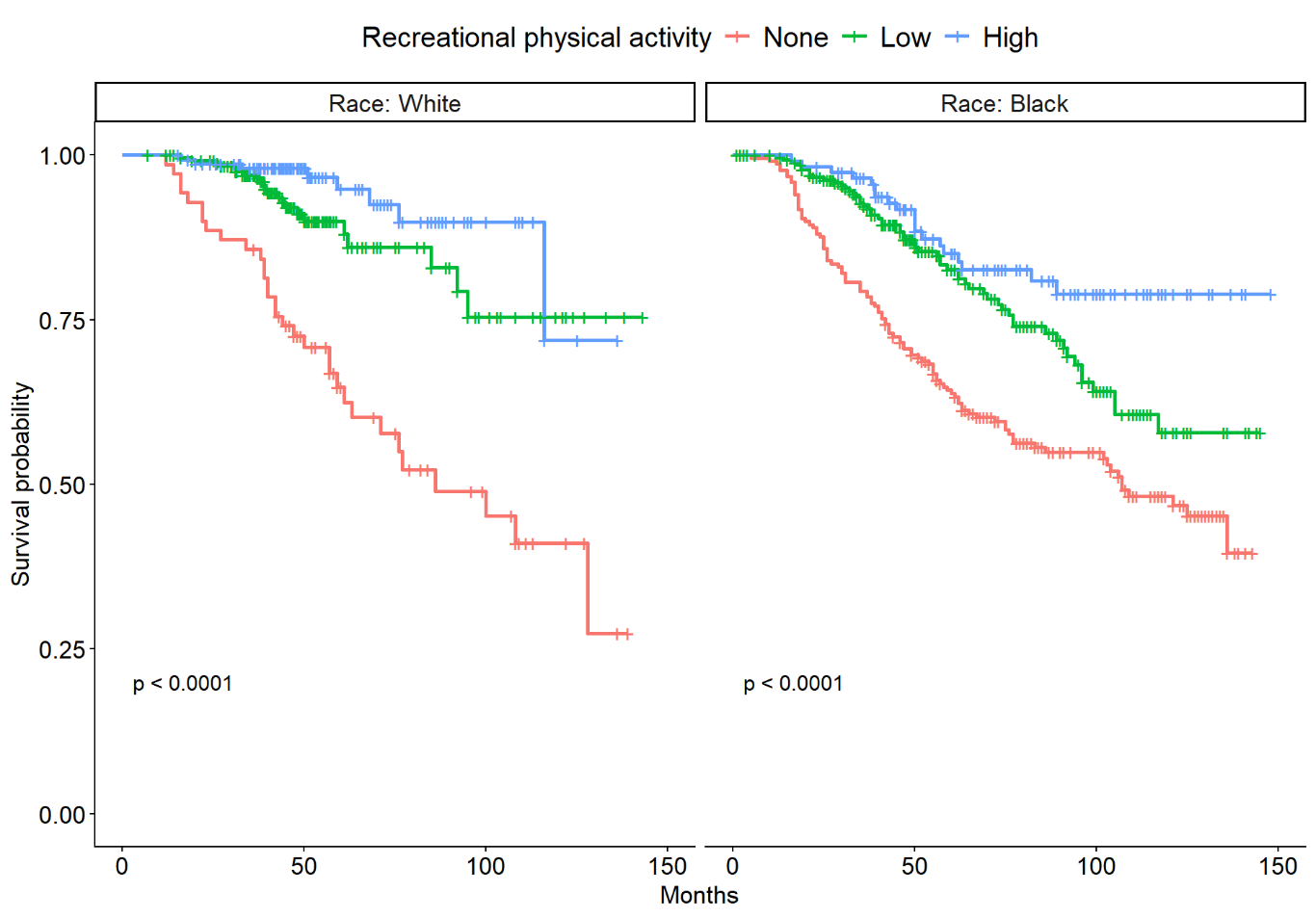
